# The Circulating Lipidome In Severe Obesity

**DOI:** 10.1101/2025.06.11.25329456

**Authors:** Mohammad Yaser Anwar, Heather M. Highland, Alexandra B. Palmer, Thy Duong, Zhaotong Lin, Wanying Zhu, Jessica Sprinkles, Daeeun Kim, Kristin L. Young, Hung-Hsin Chen, Mohanraj Krishnan, Absalon Gutierrez, Rashedeh Roshani, Elizabeth G. Frankel, Joshua Landman, Penny Gordon-Larsen, Miryoung Lee, Susan P. Fisher-Hoch, Joseph B. McCormick, Joanne Curran, John Blangero, Peter J. Meikle, Corey Giles, Jennifer E. Below, Kari E. North, Mariaelisa Graff

## Abstract

**Background:** Severe obesity (SevO; BMI ≥40 kg/m²) is rapidly increasing globally and disproportionately affects minority populations. However, it remains understudied in mechanistic and omics literature. Lipid metabolism plays a central role in obesity-related cardiometabolic disease (CMD), but the relationship between molecular lipid species and SevO is poorly understood, particularly in high-risk groups.

**Methods:** We analyzed 578 participants from the Cameron County Hispanic Cohort (CCHC) with fasting plasma lipidomic and genetic data, comparing those living with SevO (n=185) to non-obese controls (n=393). A total of 830 circulating lipid species across 49 classes were quantified. Associations between individual lipids and SevO were assessed using logistic regression and orthogonal projections to latent structures discriminant analysis (OPLS-DA). Potential causal links were assessed using network deconvolution mendelian randomization (NDMR), and influential lipids were correlated with CMD traits and DXA-derived body composition.

**Results:** Participants with SevO exhibited statistically significantly more adverse cardiometabolic risk factors than controls. Lipidomic profiling revealed broad alterations: shorter, saturated, and monounsaturated triacylglycerols were markedly elevated, while lysophospholipids, plasmalogens, cholesteryl esters, and long-chain lipids were reduced in individuals with SevO when compared to controls (BMI ≥ 18.5 and > 25 kg/m²). OPLS-DA identified over 300 influential lipid species predictive of SevO status. NDMR analyses implicated specific triacylglycerol species as potentially causally linked with SevO status. Influential lipids correlated with insulin resistance, liver steatosis, body fat measures, and HDL-C (absolute value ranges 0.2-0.4).

**Conclusions:** Our findings reveal that SevO is marked by extensive and lipid class-specific dysregulation of the circulating lipidome, with strong links to cardiometabolic risk. Notably, triacylglycerols containing shorter length acyl chains emerged as a distinctive lipid signature of SevO—consistently elevated, strongly discriminative of SevO status, and uniquely exhibiting a consistent causal relationship. Our results provide compelling evidence for a novel lipidomic pathway underpinning severe obesity and underscore critical avenues for future research into its genetic, dietary, and mechanistic determinants.

## Background

The prevalence of severe obesity (SevO, defined as BMI ≥40 kg/m²) is rapidly increasing; by 2025, at least 6% of men and 9% of women across the world are projected to live with SevO^1^. People with SevO have a reduced life expectancy compared to people with BMI in the 18-25 kg/m^2^ range due to increased risk of many diseases, including type 2 diabetes (T2D), coronary heart disease, hypertension, kidney disease, and cancer^2^. Despite SevO’s threat to public health, SevO is often an exclusion factor in epidemiological and clinical studies, limiting understanding of its mechanistic pathways in the etiology of disease. Furthermore, in understudied populations of racial and ethnic minorities, these limitations are exacerbated.

Lipid metabolism is heavily implicated in SevO^3^. Altered lipid profiles are a key differentiator of heterogeneous obesity subtypes and within-obesity variation in cardiometabolic diseases^4^. This relationship is complex as alterations in traditional/clinical lipid measures [e.g., high-density lipoprotein cholesterol (HDL-C), low-density lipoprotein cholesterol (LDL-C), and triglycerides (TRG)] are major predictors of cardiovascular disease (CVD) risk in people with SevO^5^.

These clinical measures are large macrostructures comprising 1000s of individual lipid species, each produced from interconnected metabolic pathways. As such, there is a wealth of information that may be relevant to the etiology and progression of cardiometabolic disease throughout the lipidome. These fundamental gaps in understanding the distinct structural and functional roles of lipid species in relation to SevO limit our understanding of mechanistic pathways and identification of interventional targets.

The purpose of our study was to investigate the relationship between molecular lipid species and classes with SevO in the Cameron County Hispanic Cohort (CCHC), a Mexican American population with a high burden of cardiometabolic disease^6^. Specifically, we examined associations between 830 individual lipid species (in 49 classes) and SevO status. We then used genetic association data to identify potential causal links and evaluated the clinical relevance of these influential lipids to cardiometabolic health. Our findings provide novel insights into the metabolic role and genetic underpinnings of lipid species in SevO in an understudied and high-burden population.

## METHODS

### Study population

The Cameron County Hispanic Cohort (CCHC) is a community-based, prospective study focused on self-identified Mexican Americans primarily living in Brownsville, Texas. Households were randomly selected using U.S. census tract data and a two-stage sampling method for recruitment to the cohort. So far, 5,159 individuals have been enrolled, with recruitment and 5-year follow-up visits ongoing. Extensive phenotyping was conducted at each visit, including socio-demographics, lifestyle factors, and clinical measurements collected during the initial assessment and at subsequent five-year follow-up intervals. Detailed study protocols and findings have been previously published^7^.

We used data from the earliest age when participants had their circulating lipids measured (n=2,406). We further restricted the study to those who were at least 18 years of age and had both lipid species and genotyping data collected from fasting blood samples (n = 2,105). For our study comparisons, we included only eligible participants living with severe obesity (cases) or BMI 18.5≤BMI<25 kg/m^2^ (controls), leaving a final analytic sample of 578 participants (185 classified with SevO and 393 as controls).

### Covariates, adiposity measures, and cardiometabolic disease (CMD) traits

Our assessment of cardiometabolic disease (CMD) trait associations included numerous CMD-related variables. We assessed clinical lipid measures (LDL, HDL, total cholesterol [TCH], TRG), use of lipid-lowering medications (binary; yes if ever used, no otherwise), mean fasting blood glucose (across multiple measures in the same visit, MFBG), homeostatic model assessment of insulin resistance (HOMA-IR), systolic and diastolic blood pressure (SBP & DBP), liver scans including Kappa liver stiffness test (for fibrosis) and controlled attenuation parameter [CAP]-scores (steatosis), and biomarkers of liver function (bilirubin), kidney function (creatinine), recent heart injury (troponin), and inflammation (c-reactive protein [CRP], and interleukin-6 [IL6], IL1β, IL8). In addition to anthropometry (BMI and waist circumference), we assessed body composition, measured using dual-energy X-ray absorptiometry (DXA) (Horizon W, Hologic Inc.). These measures included visceral adipose tissue (VAT), subcutaneous adipose tissue (SAT), and the VAT/SAT ratio (VSR). Total abdominal fat and VAT were measured within a 5 cm section across the abdomen, between the fourth and fifth lumbar vertebrae. SAT was estimated by subtracting VAT from the total abdominal fat in this region.

### Quantification of circulating lipid species

Fasting (overnight) blood plasma samples from each participant were used to measure circulating lipid species. More detailed information on sample collection, preparation, and preprocessing is provided in **Supplementary Section A**. Briefly, samples were processed using a protocol established by the Metabolomics Laboratory at the Baker Heart and Diabetes Institute^8^, resulting in the quantification of 830 lipid species. The results from these QC procedures are summarized in **Supplementary Section A (Table 1A)**. The 830 lipid species were analyzed individually and as members of their respective lipid classes. Class-level lipidomic measures were calculated by summing the values for individual species within each class. There were 49 lipid classes in total. The lipid species included in each class are listed in **Supplementary Section A (Table 2A)**. It should be noted that Triacylglycerol (TG) and Alkyl-diacylglycerol (TG[O]) are measured using two different techniques for the same species: single ion monitoring (SIM) for quantitation, and neutral loss (NL) ions for structural characterization. In the analysis, we tallied the species measured with each technique under separate classes for comparison.

**Table 1.**
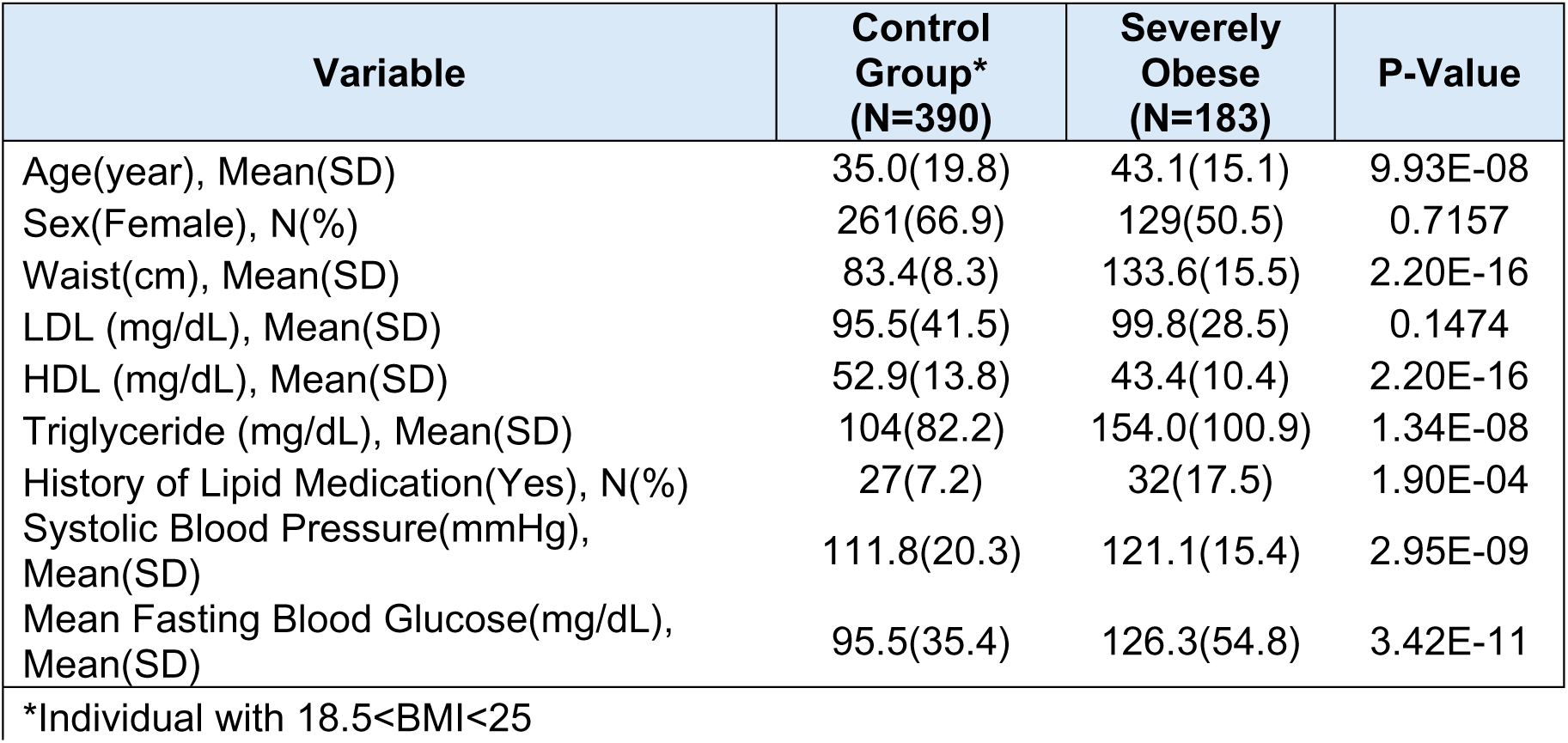
Discriptive distributions Cameron County Hispnanic Cohort subset included in the study.

| Variable | Control Group*<br>(N=390) | Severely Obese<br>(N=183) | P-Value |
| --- | --- | --- | --- |
| Age(year), Mean(SD) | 35.0(19.8) | 43.1(15.1) | 9.93E-08 |
| Sex(Female), N(%) | 261(66.9) | 129(50.5) | 0.7157 |
| Waist(cm), Mean(SD) | 83.4(8.3) | 133.6(15.5) | 2.20E-16 |
| LDL (mg/dL), Mean(SD) | 95.5(41.5) | 99.8(28.5) | 0.1474 |
| HDL (mg/dL), Mean(SD) | 52.9(13.8) | 43.4(10.4) | 2.20E-16 |
| Triglyceride (mg/dL), Mean(SD) | 104(82.2) | 154.0(100.9) | 1.34E-08 |
| History of Lipid Medication(Yes), N(%) | 27(7.2) | 32(17.5) | 1.90E-04 |
| Systolic Blood Pressure(mmHg), Mean(SD) | 111.8(20.3) | 121.1(15.4) | 2.95E-09 |
| Mean Fasting Blood Glucose(mg/dL), Mean(SD) | 95.5(35.4) | 126.3(54.8) | 3.42E-11 |

\*Individual with 18.5<BMI<25

**Table 2.**
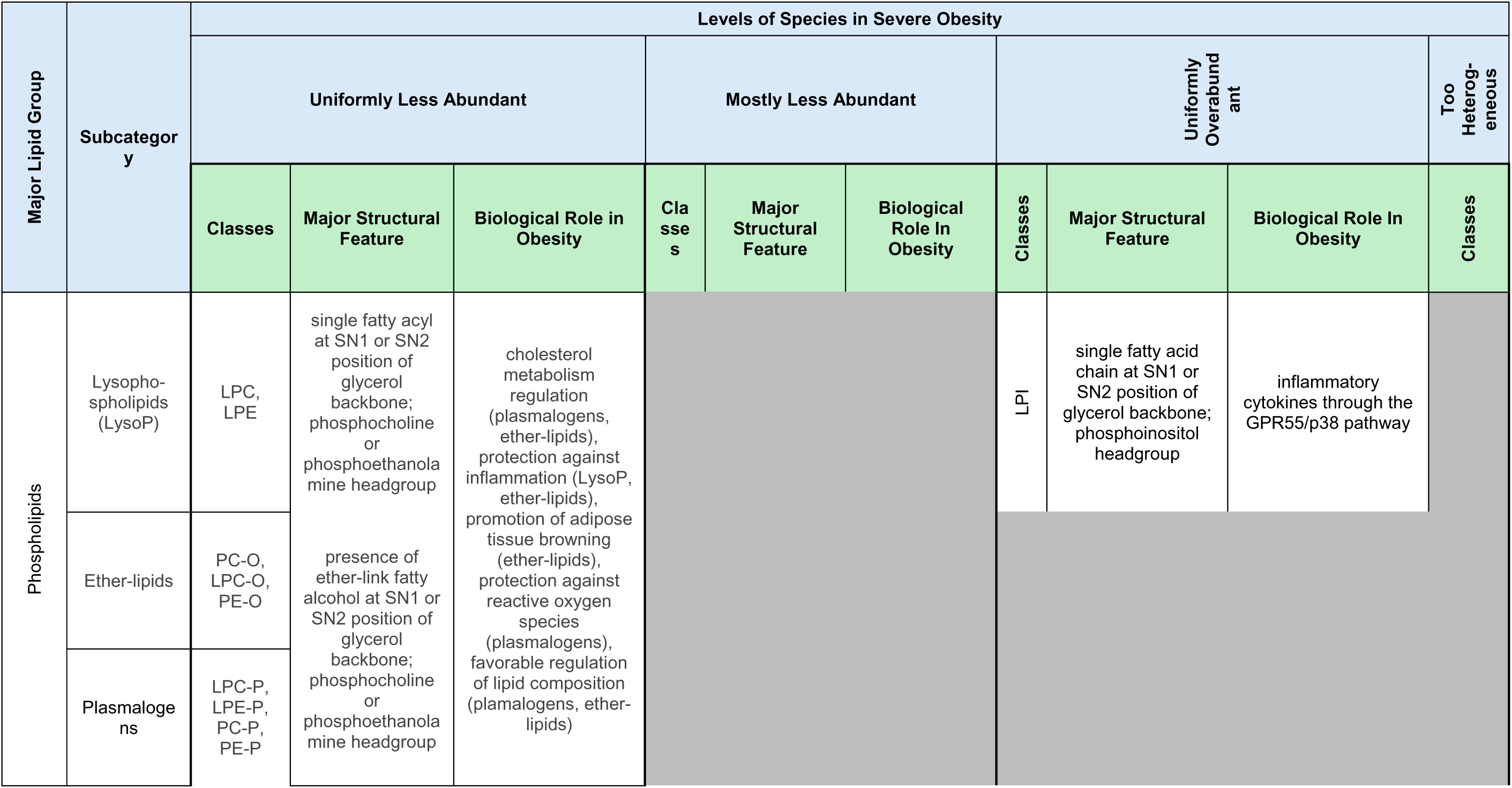
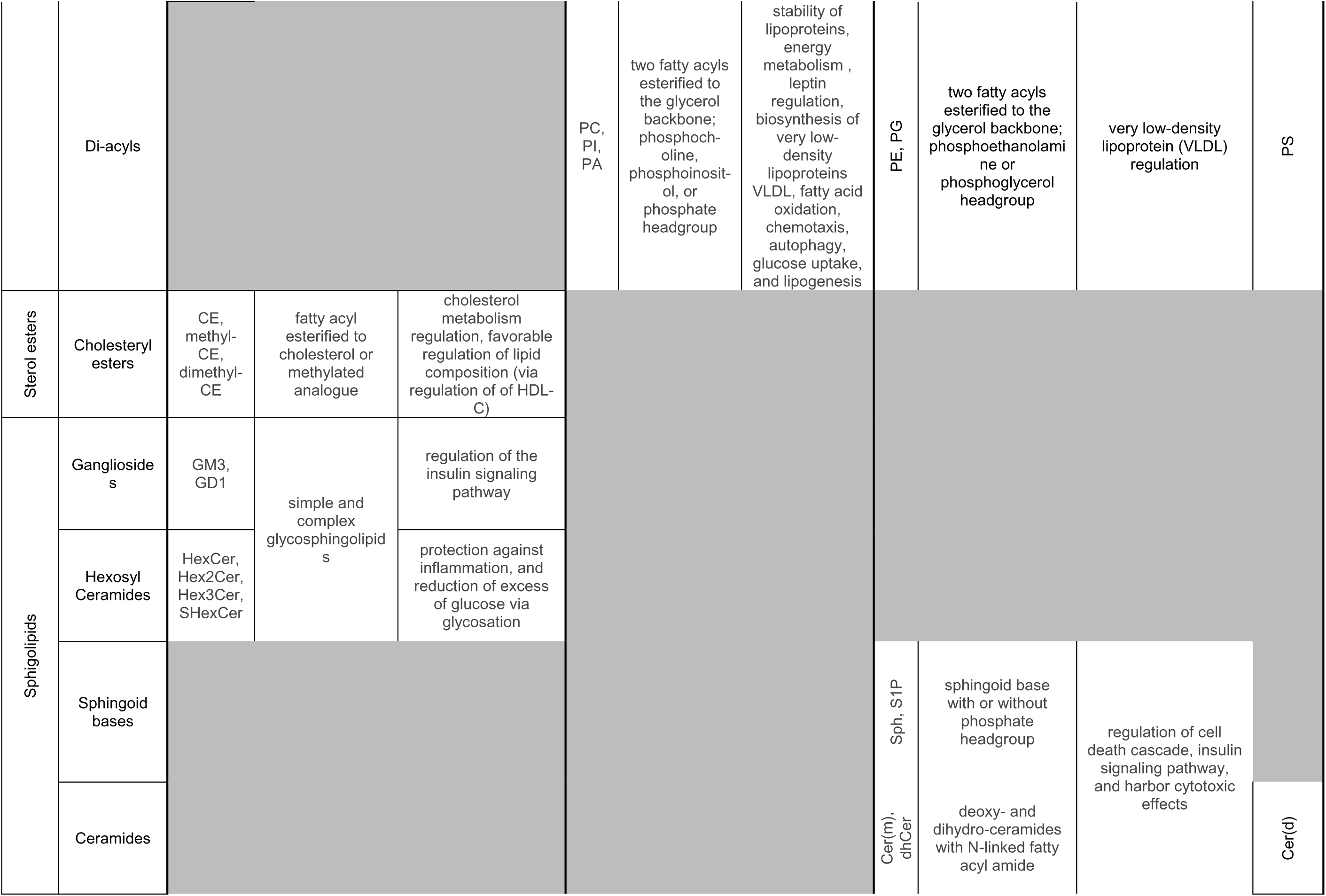
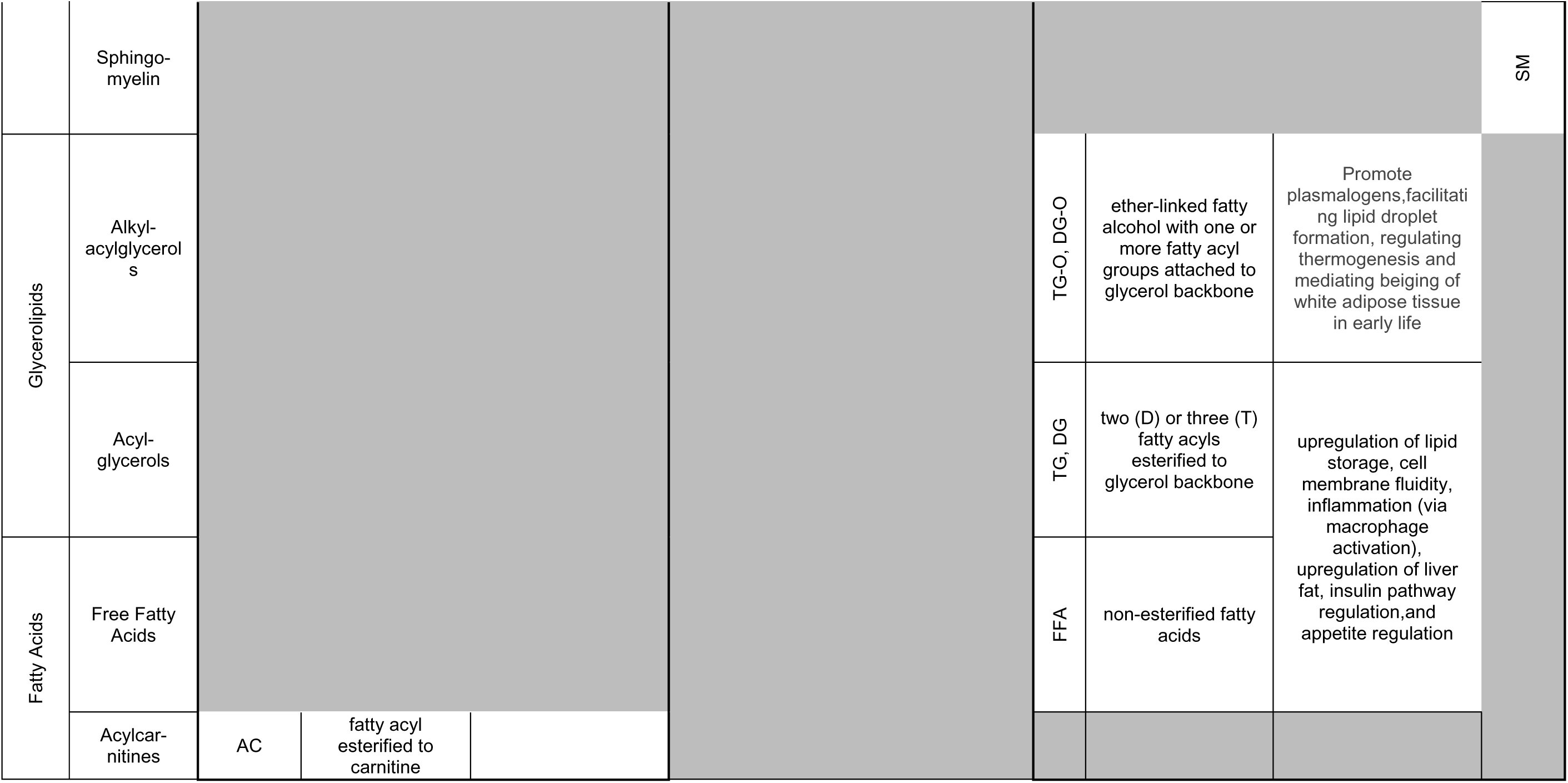
Summary of associations between severe obesity status and and major lipid groups. Results are restricted to major groups with consistent direction across all species within each class. Blackedout cells contain no character.

### Genotype array processing

Participant blood samples were genotyped on the MEGA^EX^ Array panel at the Vanderbilt University Center Genotyping core facility (VATAGE). Details on QC steps are provided in **Supplementary Section A**. Genotyped data then underwent imputation with the Trans-Omics for Precision Medicine (TOPMed) Imputation Server (freeze 8)^9^, which is an online platform developed by the University of Michigan (and can be accessed via: imputation.biodatacatalyst.nhlbi.nih.gov).

### Statistical Analysis

#### Regression modeling

We used logistic regression to examine associations between rank-normalized lipid species and lipid classes (predictors) with SevO (outcome). To account for confounding and potential non-linear effects of age, differences in age effects by sex, and ancestral confounding, regression models were adjusted for age, sex, age^2^, age×sex, age^2^×sex, and the first 4 principal components of genetic ancestry. Associations were considered significant based on Benjamini-Hochberg false discovery rate (FDR) <0.05. Given the effect of lipid-lowering medications on the lipids, we adjusted for lipid-lowering medication use (ever used vs. never used) in a sensitivity analysis to assess the robustness of regression results.

#### Discriminant Analyses

We used Orthogonal Projections to Latent Structures– Discriminant Analysis (OPLS-DA) to a) test whether the combined measured circulating lipidome distinguished severe obesity status (SevO vs. control status) and b) identify the most influential lipid species in the prediction of SevO status. OPLS-DA is a principal component analysis (PCA)-based method and a binary extension of orthogonal projections to latent structures (OPLS), a linear technique^10^. It is a particularly useful method in the context of highly collinear multivariate data, such as lipidomic profiles, which exhibit collinearity both between lipid species and across lipid classes.

We initially performed exploratory PCA analyses to evaluate the discriminative utility of lipids to distinguish SevO status and then followed with OPLS-DA models. Using ranked-normalized lipid residuals, we adjusted for the same covariates in the previously described logistic regression models, with inclusion of lipid-lowering medication use (with the latter for sensitivity evaluation). We investigated the lipidomic profiles in two different ways: 1) an overall OPLS-DA model, which included all 830 available lipid species regardless of lipid class (lipidome-wide model), and 2) separate OPLS-DA models for each lipid class, which included only the lipid species belonging to that respective class (class-specific models).

First, we assessed whether the combinations of lipid species distinguished SevO versus control status. To do this, the OPLS-DA model calculates two values: 1) R^2^Y, the total sum of variation in the outcome (Y; SevO status) explained by the model, and 2) Q^2^, an estimate of the goodness of fit for out-of-sample prediction. Higher values indicate greater variance explained and better predictive ability, respectively. We considered Q^2^ values >0.4 to be adequate^11^. We also tested the predictive performance of the lipidome-wide model by randomly splitting the data into training (70% of the sample) and validation (the remaining 30%) sets and calculating the average area under the curve (AUC) as a proportion. We reported the average value of AUC across five independent cross-validations.

Second, we identified lipid species considered to be “influential” in predicting SevO status, carrying these influential lipid species forward in subsequent analysis steps. Lipid species with variable influence on projection (VIP) scores of >1 and >1.5 were considered “influential” and “very influential”, respectively. Analyses were performed with *ropls* Bioconductor R package.

#### Examining causal associations between influential lipid species and SevO status

Since cross-sectional analysis provides limited insight into underlying causal relationships, we performed causal inference association analysis of the “influential” lipids (VIP>1) identified from our OPLS-DA models. We further restricted our analyses to lipids exhibited broad-sense heritability exceeding > 0.25, estimated using individual level data from CCHC with GCTA package (**Supplementary Section B**). Then, we employed a network deconvolution mendelian randomization (NDMR) approach, *GraphMRcML*^12^, which is an alternative to traditional MR approaches. NMDR is particularly effective when traits are collinear, associations are bidirectional, and pleiotropy is common. It infers a causal network of total effects among multiple traits and estimates the corresponding network of direct effects using a modified deconvolution algorithm. We utilized CCHC and Australia-based Busselton Health Study (BHS) genome-wide association study (GWAS) summary data for this analysis. BHS is a long-running epidemiological study of self-identified European Australians from Western Australia, and the rationale and design of the study was previously described^13^. Meta-analysis of the CCHC genetic data with BHS was intended to boost the statistical power and generalizability of causal inferencing assessments. We applied FDR correction when assessing statistical significance (FDR<0.05). Details on selecting influential lipids for NDMR and modeling steps are provided in **Supplementary Section B**.

#### Correlation between SevO-predictive lipid species and CMD traits

To determine the potential clinical relevance of our modeling findings, we assessed correlations between the OPLS-DA-identified “very influential” lipid species (VIP>1.5) and several CMD traits and measures of body composition. We first ran a series of regression models, regressing out age, sex, age^2^, age×sex, age^2^×sex, and the first 4 principal components of ancestry from lipids and CMD traits. We then used residuals from these models in a Pearson correlation analysis of CMD traits and body composition measures (vs lipids). The following CMD and body composition traits were included: systolic blood pressure (SBP), glycemic measures (HOMA-IR, MFBG, HBA1C), clinical lipids (HDL, TRG, CHOL), markers of inflammation (CRP, IL1β, IL6, IL8), adiposity (total fat, VAT, SAT, VSR), liver stiffness (kPa), and liver fat or steatosis (CAP score).

## RESULTS

### Participant characteristics

Our study design is graphically summarized in **Figure 1**. The individuals with SevO were on average 8 years older (43.1±15.1 vs. 35±19.8 in controls, p=9.93 x10^-8^), and more likely to have dyslipidemia (mean triglycerides 154±100.9 mg/dL vs 104±82.2, p=1.34×10^-8^), dysglycemia (mean MFBG 126±54.8 mg/dL vs 96±35.4, p=3.42 x10^-11^), higher blood pressure (systolic blood pressure 121±15.4 mmHg vs 112±20.3, p=2.5×10^-9^), and history of lipid medication use (32% vs 27%, p=1.9×10^-4^; **Table 1**).

**Figure 1.**
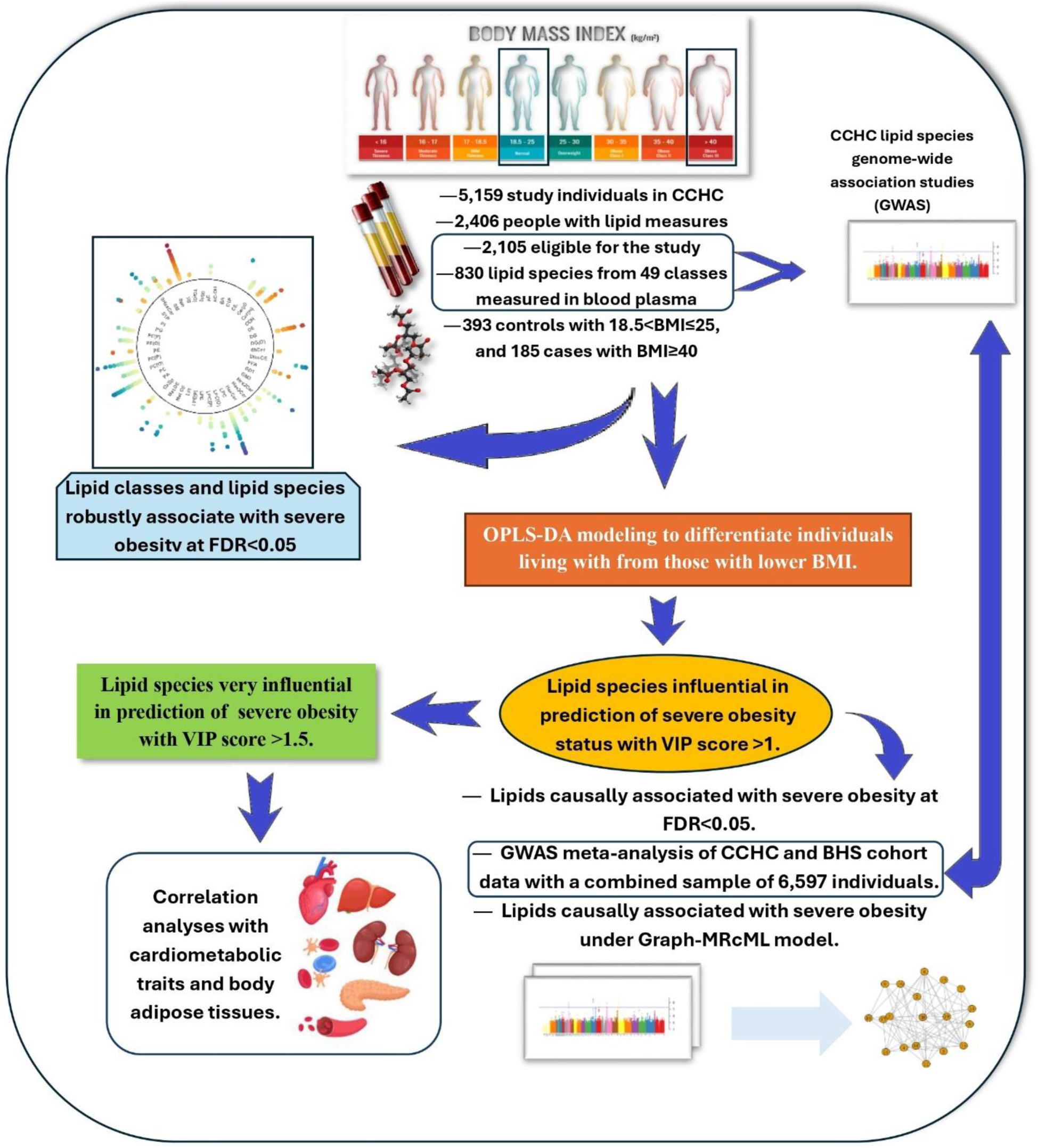
Graphical summary of the study design. Arrows indicate the direction of the study steps. VIP: variance influence on projection score, OPLS-DA: Orthogonal Projections to Latent Structures–Discriminant Analysis, NDMR: Network Deconvolution Mendelian Randomization. FDR: False Discovery Rate.

### Regression modeling identified lipid species and classes associated with SevO

In covariate-adjusted logistic regression models—bivariate association for each lipid species and class with case/control status—many species and classes differed in SevO cases compared to controls. As the volcano plot in **Figure 2a** and effect estimates in **Table S1** show, at the class level, 26 (and 11) lipid classes were significantly under-(and over-) abundant in individuals with SevO, respectively. The SevO betas (representing log-odds ratios) for the lipid classes that were overabundant ranged between 0.26 to 0.68 (OR: 1.30-1.97) for each standard deviation increase in rank-normalized lipid levels [median log-odds: 0.29 (OR: 1.34)]. In contrast, the SevO betas for the classes that were underabundant range from −0.23 to −1.34 (OR: 0.79-0.36, median −0.59 [OR: 0.55]) for each deviation increased in rank-normalized lipid levels, suggesting asymmetry in lipidomic disruption in SevO. Negative betas tended to be larger in value than positive betas. Notably, cholesteryl ester (CE) and their exogenous analogue classes, including methyl cholesteryl ester (methyl-CE), di-methyl cholesteryl ester (dimethyl-CE), and methyl-dehydrocholesteryl esters (methyl-DE), together with sulfide hexosylceramide (SHexCer), showed very low concentrations (Betas < −1.0) in individuals with SevO. Finally, the ratio of phosphatidylcholine/phosphatidylethanolamine was significantly negatively associated with SevO.

**Figure 2.**
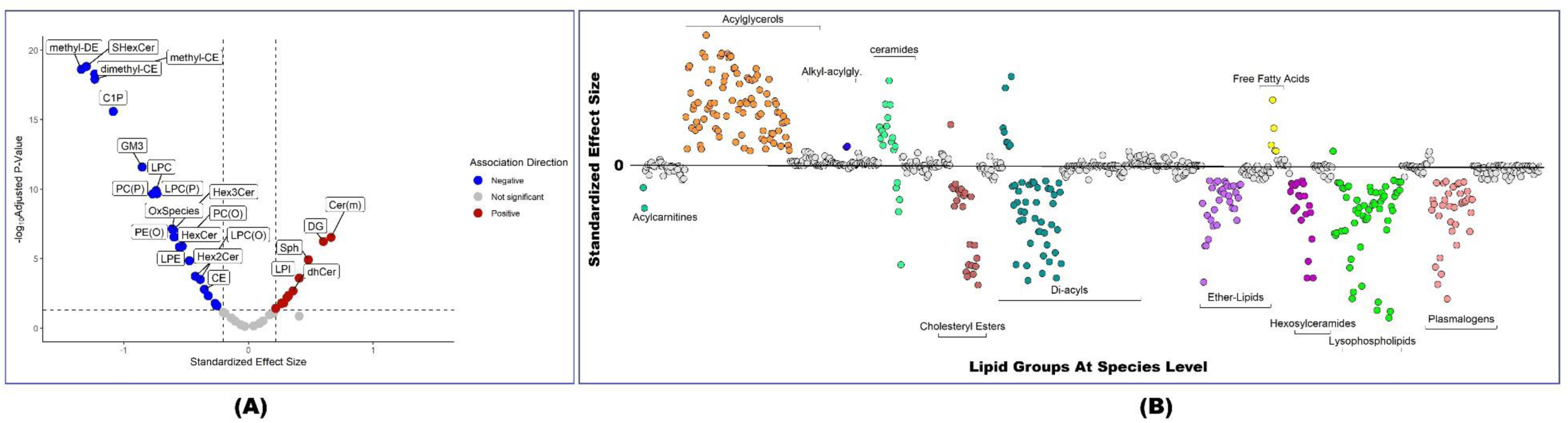
**A).** Volcano plot of associations between severe obesity status with lipids at the class level. Standardized effect sizes represent log-odds ratio for severe obesity per each standard deviation increase in rank-normalized lipid levels. Red color suggest negative association, blue represent positive association, and green demonstrate statistically insignificant association at FDR<0.05. AC=Acylcarnitine, PC(P)=Alkenylphosphatidylcholine (P), PE(P)=Alkenylphosphatidylethanolamine (P), TG(O) (NL)=Alkyldiacylglycerol (NL), TG(O) (SIM)=Alkyldiacylglycerol (SIM), DG(O)=Alkylmonoacylglycerol, PC(O)=Alkylphosphatidylcholine, PE(O)=Alkylphosphatidylethanolamine, BA=Bile acid, Cer(d)=Ceramide, C1P=Ceramide-1- phosphate, CE=Cholesteryl ester, DE=Dehydrocholesterol ester, deDE=Dehydrodemosterol ester, Cer(m)=Deoxyceramide, DG=Diacylglycerol, Hex2Cer=Dihexosylceramide, dhCer=Dihydroceramide, dimethyl-CE=Dimethyl-cholesteryl ester, COH=Free cholesterol, FFA=Free fatty acid, GD1=GD1 ganglioside, GM3=GM3 ganglioside, AC-OH=Hydroxylated acylcarnitine, LPC(P)=Lysoalkenylphosphatidylcholine (P), LPE(P)=Lysoalkenylphosphatidylethanolamine (P), LPC(O)=Lysoalkylphosphatidylcholine (LAF), LPC=Lysophosphatidylcholine, LPE=Lysophosphatidylethanolamine, LPI=Lysophosphatidylinositol, methyl-CE=Methyl-cholesteryl ester, methyl-DE=Methyl-dehydrocholesteryl ester, HexCer=Monohexosylceramide, OxSpecies=Oxidised lipids, -=PC/PE Ratio, PA=Phosphatidic acid, PC=Phosphatidylcholine, PE=Phosphatidylethanolamine, PG=Phosphatidylglycerol, PI=Phosphatidylinositol, PIP1=Phosphatidylinositol monophosphate, PS=Phosphatidylserine, SM=Sphingomyelin, Sph=Sphingosine, S1P=Sphingosine-1-phosphate, SHexCer=Sulfatide, TG (NL)=Triacylglycerol (NL), TG (SIM)=Triacylglycerol (SIM), Hex3Cer=Trihexosylcermide, ubiquinone=Ubiquinone. **B).** Associations between severe obesity status with lipids at the specie level. Colors represent distinct lipid groups, and each dot represent a specie within each class. If association was not significant at False discovery level (<0.05), association is represented with gray colored dot.

At the species level, 362 of the 830 lipid species were underabundant, and 222 lipid species were overabundant in SevO versus control status (**Table S2**). There were multiple significant patterns at the species level. For underabundant species, we observed strong negative associations (Beta < −0.5 [OR: 0.61]) for very long chain saturated lysophosphatidylcholine (LPC) species, which suggests an underabundance in individuals living with SevO. These are complemented by a variety of other lysophosphatidylcholine species, including LPC 18:2, LPC 18:1, and several odd-chain LPC species (19:0, 17:0, 15-MHDA, 17:1, 19:1). Amongst the strongest negative associations are very long chain ether-linked lysoalkylphosphatidylcholine species [i.e. LPC(O-22:0), LPC(O-22:1), LPC(O-24:0), LPC(O-24:2), LPC(P-20:0), LPC(P-18:0), LPC(O-24:2), etc.] Although these are structurally similar to lysophosphatidylcholine species, their biosynthetic pathway relies on different mechanisms.

Like class-level associations, most cholesteryl esters were significantly underabundant in SevO, with some neutral. However, two CE species show strong positive associations: CE(16:1) and CE(20:3). In addition, lipid species containing 22:6 (DHA, docosahexaenoic acid), a type of omega-3 fat, were significantly underabundant in SevO [e.g., PC(18:1_22:6) (a), PE(O-18:1/22:6), etc.] Similarly, species containing a polyunsaturated linoleic acid (18:2) acyl chains were generally underabundant in SevO. However, species containing 20:4 (arachidonic acid) acyl chains were variably associated with SevO status and at lower magnitudes.

Overall, the most overabundant species were triacylglycerols. However, the most overabundant species (in SevO group compared to controls) tended to contain shorter and more mono-unsaturated or saturated fatty acyls [i.e., TG(48:1) [NL-16:1], TG(48:0) [NL-16:0], TG(50:1) [NL-16:0], TG(49:1) [NL-17:1], TG(49:1) [NL-16:1], etc.] Similar pattern of significant overabundance of diacylglycerol species was observed complementary to triacylglycerols [e.g., DG(16:0_16:1) and DG(16:0_16:0)]. In contrast, the tri-/diacylglycerol species with higher total numbers of carbons, or more double bonds, trend towards no association. In fact, there was a trend for positive associations with species containing 16:0 and 16:1 (not ubiquitous in all classes, but present in many classes): PI(16:0_16:1), PE(16:0_16:1), FA(16:1), FA(16:0), CE(16:1), DG(16:0_16:1), TG(48:1) [NL-16:1], and TG(48:0) [NL-16:0].

### Lipid class groupings based on uniformity of their constituent lipid species (uniformly underabundant vs. overabundant in SevO)

To summarize the lipid species association analyses, we categorized lipid classes into groups based on the direction and statistical significance of their respective lipid species results: uniformly and less abundant in SevO, mostly less abundant in SevO, uniformly overabundant in SevO, and too heterogeneous to categorize SevO (**Table 2; Figure 2b**).

We identified 19 lipid classes whose lipid species were uniformly underabundant in people with SevO. These 19 classes had four underlying molecular structures (**Table 2**): 1) phospholipids, including lysophospholipids, ether-lipids, and plasmalogens, 2) sterol esters, including cholesteryl esters and related species, 3) sphingolipids, including ganglioside species and hexosylceramide species, and (4) fatty-acid-based acylcarnitines (AC). We additionally identified three phospholipid classes (PC, PI, and PA, belonging to the Di-acyl sub-group) (**Table 2**), whose lipid species were generally (though not uniformly) less abundant in people with SevO (**Figure 2b**; **Table S2**).

We also identified 12 lipid classes whose constituent lipid species were uniformly overabundant in people with SevO. Ten of these 12 lipid classes belonged to three broad categories: 1) glycerolipids including di- and tri-acylglycerols and alkylglycerols, 1) deoxy and dihydroceramides, and sphingoid precursors of ceramids, and 3) di-acyl phospholipids including phosphatidylethanolamines and phosphatidylglycerols (**Table S1**; **Table 2**; **Figure 2b**).

Lipid species within the remaining classes (ceramides, sphingomyelins, and phosphtidylserines), showed inconsistent, or heterogenous, associations with SevO status. In the sensitivity analysis adjusting for lipid-lowering medication use, associations between SevO status and cholesteryl ester species were attenuated (**Figure S1**), whereas associations with acylglycerols and deoxyceramides were strengthened.

### OPLS-DA modeling differentiates participants with SevO from controls

We next assessed the predictive power of the full lipidome (consisting of 830 lipid species) to distinguish participants with SevO from controls. The initial exploratory Principal Component Analysis (PCA), to reduce dimensionality of the lipidomic data, showed that the lipidome significantly differentiated participants with SevO from controls (**Figures S2A-2D**). The lipidome-wide OPLS-DA model revealed a clear separation between participants with SevO and controls based on their lipidomic profiles. The total Q^2^ was 0.558, which surpasses the minimum threshold of 0.4, thus indicating adequate discriminative ability of the model. Further, a large proportion of the variation in SevO status was explained by the lipidome (*R^2^*Y=0.638, which suggests negligible overfitting given the proximity to Q2 value) (**Table 3, Figure 3A, Figure S3A-C).** Internal validation of the model demonstrated strong predictive performance, with a mean area under curve (AUC) of 0.90 (across five different cross-validations), indicating excellent model accuracy. In sensitivity analysis, the OPLS-DA model adjusted for lipid-lowering medication utilization history performed similarly well, though the internal validation yielded a slightly lower AUC (0.86) (**Table S3**).

**Figure 3.**
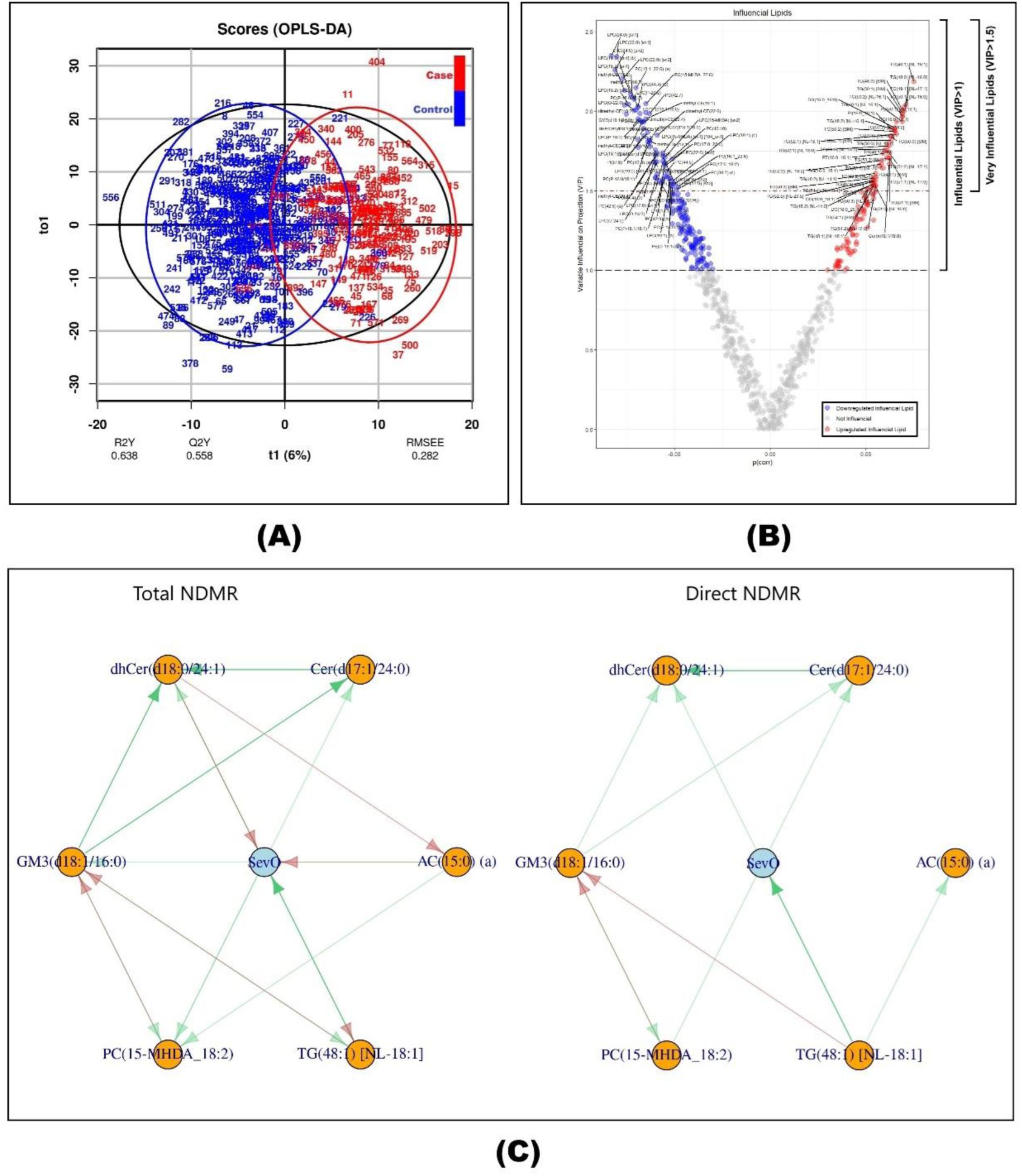
**A).** Prediction model based on the orthogonal projections to latent structures discriminant analysis (OPLS-DA) model. Red color (1) indicate severely obese, and blue (0) control individuals. Numbers represent individuals and assigned randomly (substituting study-specific IDs). **t1 & t01** represent orthogonal components (modified principal components), and axes represent orthogonal scores of across group variation (t1), and within group variation (to1). Percentages represent proportion of variation captured by the given orthogonal component. **R2Y(cum):** Is the total sum of variation in Y explained by the model. **Q2Y**: estimation of predictive performance in proportion. A threshold 0.5 is generally used as a cut-point for goodness of prediction. RMSEE: The square root of the mean error between the actual and the predicted responses. **B).** Volcano Plot of the lipids that are influential in prediction of severe obesity (with variable influence on projection >1). AC=Acylcarnitine, PC(P)=Alkenylphosphatidylcholine (P), PE(P)=Alkenylphosphatidylethanolamine (P), TG(O) (NL)=Alkyldiacylglycerol (NL), TG(O) (SIM)=Alkyldiacylglycerol (SIM), DG(O)=Alkylmonoacylglycerol, PC(O)=Alkylphosphatidylcholine, PE(O)=Alkylphosphatidylethanolamine, BA=Bile acid, Cer(d)=Ceramide, C1P=Ceramide-1-phosphate, CE=Cholesteryl ester, DE=Dehydrocholesterol ester, deDE=Dehydrodemosterol ester, Cer(m)=Deoxyceramide, DG=Diacylglycerol, Hex2Cer=Dihexosylceramide, dhCer=Dihydroceramide, dimethyl-CE=Dimethyl-cholesteryl ester, COH=Free cholesterol, FFA=Free fatty acid, GD1=GD1 ganglioside, GM3=GM3 ganglioside, AC-OH=Hydroxylated acylcarnitine, LPC(P)=Lysoalkenylphosphatidylcholine (P), LPE(P)=Lysoalkenylphosphatidylethanolamine (P), LPC(O)=Lysoalkylphosphatidylcholine (LAF), LPC=Lysophosphatidylcholine, LPE=Lysophosphatidylethanolamine, LPI=Lysophosphatidylinositol, methyl-CE=Methyl-cholesteryl ester, methyl-DE=Methyl-dehydrocholesteryl ester, HexCer=Monohexosylceramide, OxSpecies=Oxidised lipids, -=PC/PE Ratio, PA=Phosphatidic acid, PC=Phosphatidylcholine, PE=Phosphatidylethanolamine, PG=Phosphatidylglycerol, PI=Phosphatidylinositol, PIP1=Phosphatidylinositol monophosphate, PS=Phosphatidylserine, SM=Sphingomyelin, Sph=Sphingosine, S1P=Sphingosine-1-phosphate, SHexCer=Sulfatide, TG (NL)=Triacylglycerol (NL), TG (SIM)=Triacylglycerol (SIM), Hex3Cer=Trihexosylcermide, ubiquinone=Ubiquinone. **C).** Total and direct network deconvolution mendelian randomization (NDMR) model. Green colors represent positive and red colors denote negative causal association, while arrows represent causal direction. Intense colors indicate significant associations at FDR<0.05 level. Only TG specie retain significance (as SevO causing specie) in both total and direct models. AC: acylcarnitine, GM3: GM3 ganglioside, dhCer: Dihydroceramide, PC: Phosphatidylcholine, Cer(d): Ceramide, SevO: Severe Obesity, TG: Triacyglycerol.

**Table 3.**
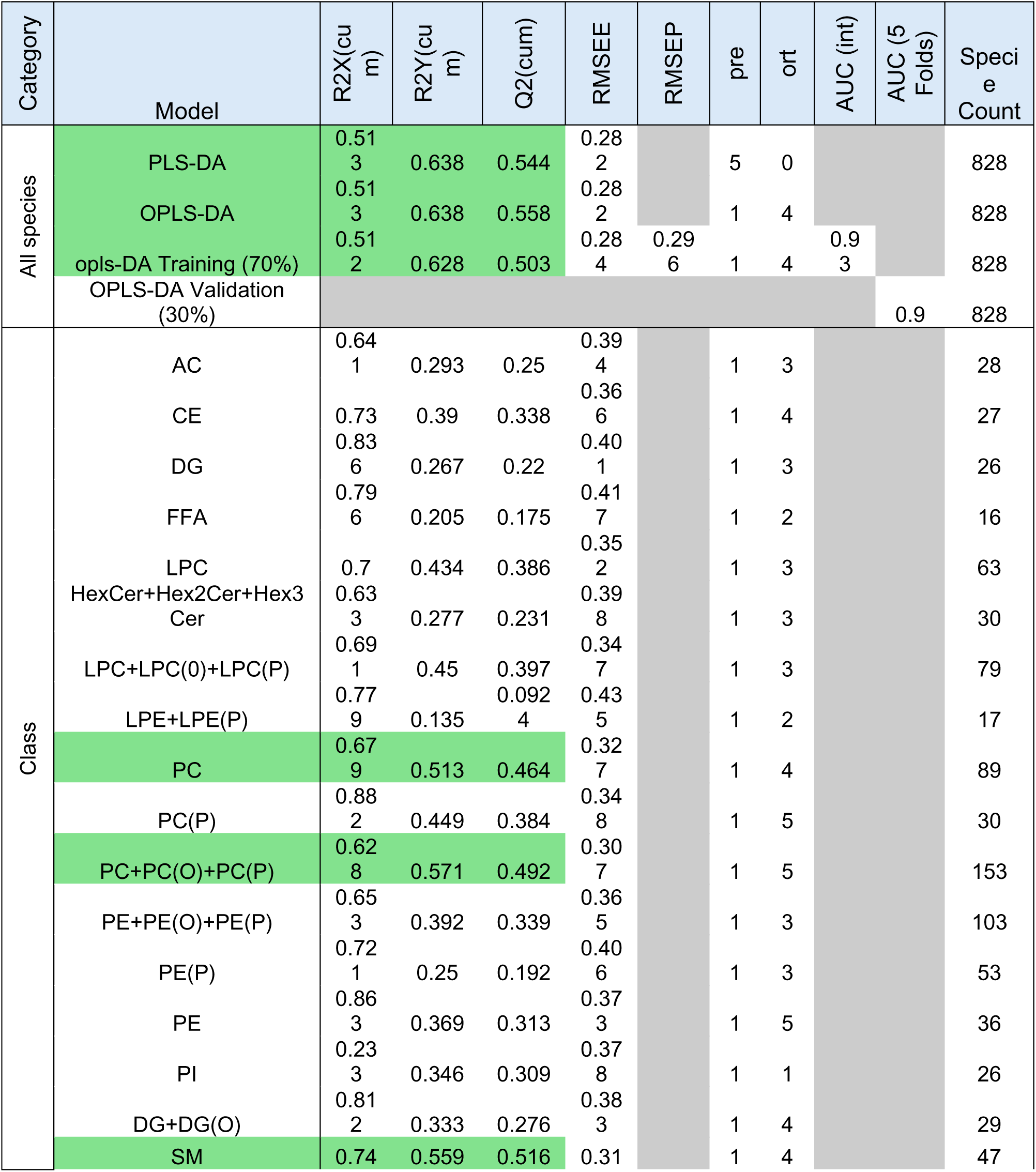
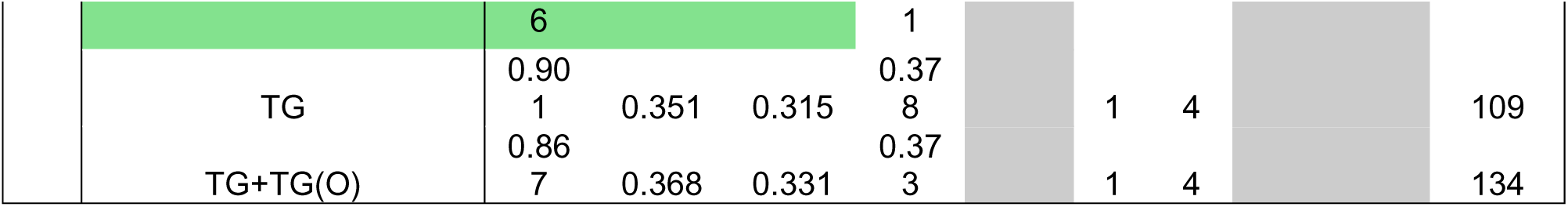
Differentiatory performance of lipid species to predict severe obesity status. Both OPLS-Da and OPLS models suggested excellent perdictive performance (Q2>0.4 and R2Y>0.5), and interval validation generated area under curve (AUC)=0.91. **R2X(cum)**: Is the sum of predictive + orthogonal variation in X that is explained by the model, **R2Y(cum)**: Is the total sum of variation in Y explained by the model, **RMSEE**: The square root of the mean error between the actual and the predicted responses, **RMSEP**: The square root of the mean error between the actual and the predicted responses in test set, **ort**: orthogonal elements, **pre**: predictive PCs [principal components] used.

We then used class-specific OPLS-DA models to evaluate the ability of each of the 49 different lipid classes (plus two simulated TG classes) to distinguish cases and controls. The following class-specific OPLS-DA models demonstrated adequate predictive ability to distinguish participants with SevO: phosphatidylcholine class (Q^2^ = 0.464, R^2^Y = 0.513), combined phosphatidylcholine species + phosphatidylcholine derivatives classes (Q^2^ = 0.492, R^2^Y = 0.517), and sphingomyelin class (Q^2^ = 0.516, R^2^Y = 0.559). (**Table 3**)

A total of 300 lipid species were identified as “influential” predictors (VIP >1) and 84 were “very influential” predictors (VIP>1.5) of SevO in the lipidome-wide OPLS-DA model (**Figure 3A**). Of the 300 species in the “influential” subset, 111 were overabundant and 189 were underabundant in individuals with SevO. This finding remained largely robust, even after adjusting for lipid-lowering medication (**Table S4**).

Among the 111 overabundant lipid species, most (71) were acylglycerols and deoxyceramides (**Figure 3B**, **Table S4**). The top overabundant influential lipids were overwhelmingly saturated or mono-unsaturated acylglycerols with shorter length acyl chains that were the most overabundant species in regression analyses (see **Table S2, Table S5**). While the top underabundant influential species exhibited greater diversity, lysophospholipids, phosphocholines, and cholesteryl ester species comprised the largest groups (representing 83 of the 189 identified underabundant species). Most of these species are lysophosphocholines or contain longer length acyl chains.

### Causal Inference: Triacylglycerol species estimated to be causally linked to SevO

SNP-based heritability was estimated using individual-level data. We found that 172 of the 300 influential lipid species (VIP>1) had at least moderate broad-sense heritability (*H_2_*>0.25). The median heritability for the entire lipid set, irrespective of association with SevO, was 37% (**Figure S4a-b, Table S6**). After FDR correction, we identified 31 lipid species (spanning 13 different classes) significantly associated with SevO in forward or reverse directions using inverse variance weighted MR method (FDR<0.05) (**Supplementary Section B**), and therefore eligible for causality assessment under *GraphMRcML* framework. After excluding genetically correlated species (*r_g_*<0.8), six species, each representing a distinct lipid class—including dehyroceramide, ceramide, GM3 ganglioside, acylcarnitine, phosphatidylcholine, and triacylglycerol—remained and were included in the NDMR model. After adjusting for pleiotropy, mediation, and bidirectionality within the NDMR framework, the total effect model (Figure 3C) showed that species representing dihydroceramide and acylcarnitine classes were negatively causal to SevO status (red arrows), and the species representing the TG class was positively causal to SevO status (green arrow). Concurrently, the SevO status was a positive regulator of species representing GM3 ganglioside, ceramide, and dihydroceramide classes, which suggested that the association with the dihydroceramide species was bidirectional. Contrastingly, in the direct effect model, SevO status was only positively causally related to the dihydroceramide species, while SevO status was causally affected by the triacylglycerol species. However, after adjusting for multiple testing, only the lipid species representing triacylglycerol class remained the sole positive regulator of SevO status at FDR<0.05 in both total and direct effect models. (**Figure 3C**).

### Clinical correlations and pathophysiologic implications: Very influential lipids predictive of SevO status were correlated with cardiometabolic disease (CMD) traits

Finally, we assessed the correlation between “very influential” lipid species in SevO prediction (84 species with VIP>1.5) with CMD traits (**Figure 4**). This subset of very influential species represents seven distinct classes. Correlation between select lipid species and examined traits ranged from −0.4 to 0.4. Very influential di- and tri-acylglycerols, which are rich in saturated and monounsaturated fatty-acid chains, were positively correlated with liver steatosis score (CAP), glucose traits (FSBG and HOMA-IR), and measures of body adipose tissues (specifically VAT), but negatively correlated with HDL-C. It was noted that lower chain triacylglycerols were relatively more positively correlated with HOMA-IR, VAT, and CAP than longer chain species [e.g., comparing TG(48:0) versus TG(54:1), etc.] However, higher chain acylglycerols were (relatively) more positively correlated with total cholesterol and more negatively correlated with HDL. In contrast, very influential phosphatidylcholine, lysophosphocholines, alkenylphosphatidylcholine, methyl-cholesteryl ester, and sulfatide hexocylceramides were negatively correlated with glucose traits, body adipose tissue measurements, and liver fat scores (CAP) but positively correlated with HDL-C and LDL-C. A more detailed inquiry showed that except for plasmalogens, very influential species in other classes containing shorter 16-22 carbon acyl chains were more strongly positively correlated with total cholesterol. In contrast, very long-chain lysophosphatidylcholines were more positively correlated with HDL-C. Also, lysophosphatidylcholines containing shorter carbon chains were more negatively correlated with CRP. Very influential plasmalogens were strongly positively correlated with both total cholesterol and HDL-C. Multiple very influential lipids in phosphatidylcholine classes were also negatively correlated with troponin, a marker of recent cardiac injury. Interestingly, none of the very influential lipids in any class strongly correlated with VSR or the visceral to subcutaneous adipose tissue ratio.

**Figure 4.**
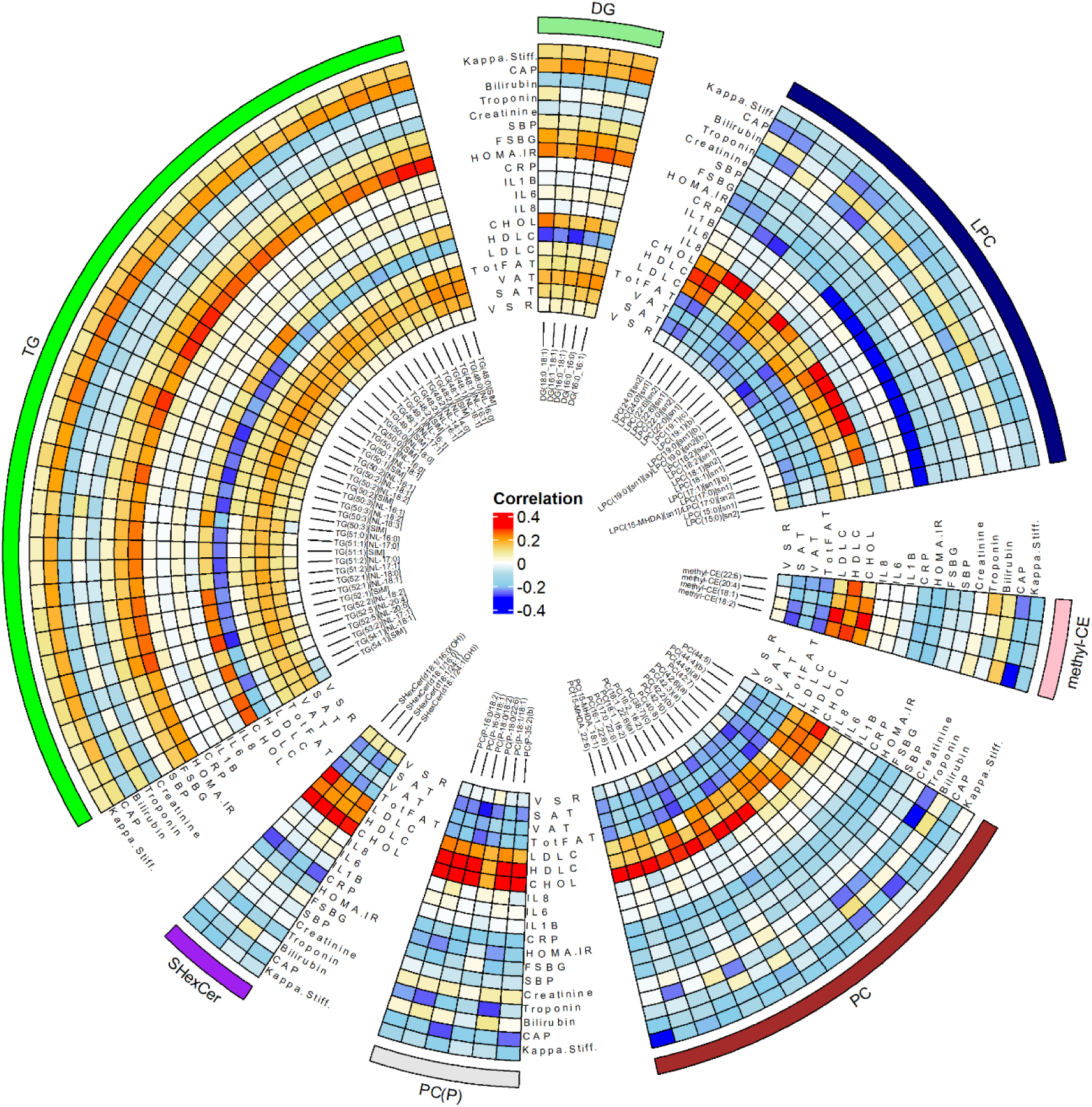
Adjusted Pearson’s correlation between 84 very influential lipids (in prediction of severe obesity, with variable influence on project score >1.5), and metabolic traits in CCHC population. Blue colors suggest negative correlation, while red colors indicate positive correlations. Color intensity indicates degree of correlation in percentage. The outermost color strip represent distinct lipid classes. CCHC: Cameron County Hispanic Cohort. DG=diacylglycerol, DG=triacylglycerol, LPC=Lysophosphatidylcholines, PC=phosphatidylcholines, methyl-CE=methylat cholesteryl esters, SHexCer=sulfatide hexocyl ceramides, PC(P)= Alkenylphosphatidylcholine, Kapp.Stiff.=Kappa Stiffness, CAP=controlled attenuation parameter, SBP=Systolic Blood Pressure, FSBG=Fasting Blood Glucose, HOMA.IR=Homeostatic Module Assessment Insulin Resistance, IL1B=Interleukin 1Beta, CHOL=Cholesterol, HDLC=High-density Lipoprotein Cholesterol, LDLC= Low-density Lipoprotein Cholesterol, TotFat= Total Body Fat, VAT= Visceral Adipose Tissue, SAT=Subcutaneous Adipose Tissue, VSR= VAT to SAT Ratio.

An overall summary of observed patterns in our study is shown in **Figure 5**. In general, lipids containing shorter, saturated, and monounsaturated acyl chains were more likely to be overabundant in SevO, particularly acylglycerols. In contrast, lipids containing long and very long acyl chains were primarily underabundant in SevO status, encompassing wider lipid groups including lysophospholipids, plasmalogens, ether-lipids, hexosylceramides, gangliosides, and sterol esters. The ceramide pool was likely altered, where deoxy- and dihydroceramides were overabundant. The collective lipidome was a significant predictor of SevO status. At the species level, acylglycerols, lysophospholipids, sterol esters, and phosphocholines were highly influential in predicting SevO status. These highly influential lipids were strongly correlated with multiple metabolic traits, including HDL-C, liver stiffness, insulin resistance, and inflammation markers.

**Figure 5.**
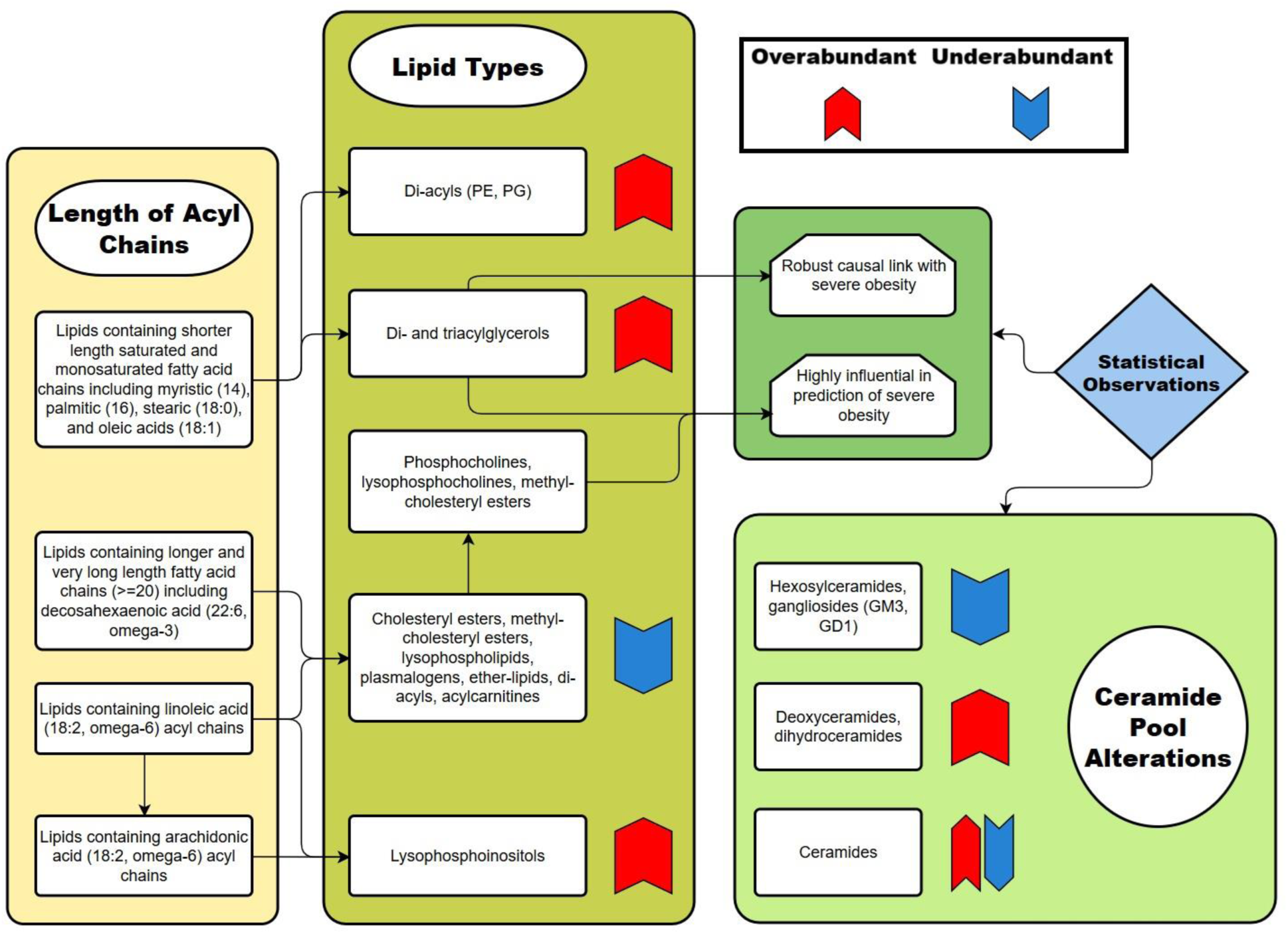
Diagram plot summarizes the study findings. Arrows indicate increased or decreased levels in severe obesity status. Colored arrows represent association direction with severe obesity. Diagram arrows are used to link observations.

## DISCUSSION

Our findings demonstrate that SevO is characterized by widespread, class-specific perturbations of the circulating lipidome, many of which are linked to established cardiometabolic risk factors^14^. Among these, triacylglycerols containing saturated and monounsaturated shorter length acyl chains emerged as a distinctive lipid signature of SevO—consistently elevated, highly differentiative of SevO status, and uniquely demonstrating a robust and consistent causal association. Additional lipid species, including ceramides, lipoproteins, acylcarnitines, phosphatidylcholines, and cholesteryl esters, also contributed to the differentiation of SevO, albeit with varying degrees of consistency. In contrast, lipids with longer (generally ≥ 18 carbons) and very-long-length (≥ 21 carbons) acyl chains were generally underabundant in individuals with SevO, potentially reflecting the influence of population-specific nutritional and genetic factors. Together, these results offer novel mechanistic insights into the lipidomic underpinnings of severe disease and provide a foundation for future translational and therapeutic investigation.

We found that acylglycerols (mainly di- and triacylglycerols) were consistently overabundant in participants with SevO, were influential or highly influential in distinguishing SevO status from the control group, and were the only group with a consistent causal regulatory effect on SevO. While precise molecular characterization of the roles of all individual acylglycerols in SevO is beyond the scope of this study, their associations with SevO may be bidirectional in nature. Excess energy intake is primarily stored as triglycerides in the body^15^. Increased triglycerides in adipose tissue trigger the reduction of HDL-C via cholesteryl ester transfer protein (CETP) activity^16^, dysregulation of triacylglycerol synthesis, and disruption of triacylglycerol lipolysis (mainly through insulin-dependent pathways^17^). All of these changes have been suggested to induce pathologic changes such as lipodystrophy and elevated steatosis^18^ that precede morbid obesity. Triacylglycerols also promote macrophage activation^19^, which induces or exacerbates inflammation. Furthermore, di- and triacylglycerols are known appetite regulators^20^ and, under certain pathologic contexts, exacerbate insulin resistance^21^. This creates a vicious cycle in which insulin resistance promotes further adiposity and the overproduction and release of free fatty acids from adipose tissue (we observed a corroborating overabundance of increased free fatty acids in SevO).

Equally notable, the most overabundant and highly influential acylglycerols contained shorter saturated or monounsaturated acyl chains (e.g., 14-18 carbons). This pattern of overabundant shorter length saturated- and monounsaturated-containing species was consistent across many other classes, though not as widespread or significant as in acylglycerols. The saturated myristic acid (14:0) is a potent inducer of adipose tissue inflammation^22^ and is highly atherogenic. The saturated palmitic acid (16:0 carbons) stimulate inflammation through reactive oxygen species (ROS) in a TLR-independent manner^23^. That is, ROS activates IL1-β, resulting in the downregulation of insulin signaling^24^. Palmitic acid is also reported to dysregulate glucose metabolism through the GPRs/NF-κB/KLF7 signaling pathway^25^. The saturated stearic acid (18:0) is inflammatory (via endoplasmic reticulum stress–mediated apoptosis^26^) and has been shown to promote food intake by reducing serum leptin through the JAK2/STAT3 pathway^27^. Generally, higher length oleic (18:1) and stearic acid are less atherogenic than palmitic acid^28^.

While triacylglycerols had the most consistent relationship with SevO, several other lipid species were overabundant in people with SevO compared to those with lower BMI. Notably, sphingosines, deoxyceramides, and dihydroceramides were consistently overabundant within the sphingolipid category. The overabundance of sphingosines and dihydroceramides in SevO may implicate an increased synthesis of sphingolipids, which are precursors to ceramides. The concurrent negative associations of ganglioside GM3 and ceramide 1-phosphate species with SevO status provide additional evidence for this “altered ceramide profile” proposition. It should be noted that dihydroceramides are linked to increased adipogenesis in mice^29^. The overabundance of noncanonical deoxyceramides may be the result of serine palmitoyltransferase (SPT) activity, where, under metabolic stress, SPT incorporates L-alanine or glycine in the place of serine to produce deoxysphingoid base. These altered compounds have lipotoxic effects and are cytotoxic, largely attributed to the inability to undergo further metabolization, like other sphingolipids.

Additionally, we found that lysophosphoinositol species were consistently overabundant in individuals living with SevO. Our observation largely corroborates published observations showing that lysophosphoinositols induce inflammatory cytokines via the GPR55/p38 pathway, were positively correlated with insulin secretion, and were suggested as early biomarkers of prediabetes^30^.

Cholesteryl esters and their plant-based analogues, including methyl-cholesteryl and dimethyl-cholesteryl esters, were significantly underabundant in SevO. Cholesteryl esters are transferred from HDL particles to apolipoprotein B (apoB) by cholesteryl-ester transfer protein (CETP) in exchange for triglycerides^16^. Our observed underabundance likely reflects elevated CETP activity and decreased HDL-C levels^31^. It should be noted that commonly used lipid-lowering medications can potentially induce such a decrease, given that they act primarily on cholesterol^32^ (i.e., **Figure S1**). Therefore, the significant underabundance of cholesteryl esters and their plant-based sterol analogues (i.e., methyl and di-methyl esters) may reflect population-specific dietary patterns, medication use or genetic underpinnings, important factors to be considered in future research.

We also observed a consistent underabundance of hexosylceramides in individuals with SevO, which further corroborates the altered ‘ceramide pool’^33^ proposition. Altered glycoceramides may occur from decreased synthesis or increased degradation. In such a milieu, the concurrent underabundance of hexosylceramides and increased level of pro-inflammatory ceramides likely exacerbate the cascade of pathophysiological effects (mainly apoptosis) that otherwise would be inhibited by hexosylceramides^34^.

Phospholipids including ether-lipids, lyosphospholipids, and plasmalogens were also underabundant in SevO. The lower lysophospholipid levels observed in participants living with SevO have been experimentally demonstrated to increase the risk of inflammation, insulin resistance, and fatty liver^35^. The significant underabundance of DHA-containing lipids further corroborates this observation. Anti-inflammatory properties of ether-lipids are conveyed through modulation of the PI3K/Akt/mTOR and RAS/RAF/MEK/ERK pathways^36^. Ether-lipids alter cholesterol metabolism by interfering with its transport from the plasma membrane to the endoplasmic reticulum^37^. Plasmalogens, a subset of ether-lipids, on the other hand, are plasma membrane constituents that facilitate increased resistance to oxidative stress and protect against reactive oxygen species^38^. Plasmalogens are also important regulators of cholesterol biosynthesis, contribute to intracellular cholesterol transport, and facilitate HDL-mediated cholesterol efflux^39^ and mitochondrial fission in brown adipose tissue^40^. The negative association with SevO status in our study corroborates previous observations where dysregulation of peroxisomal membrane protein Pex11a, involved in the synthesis of plasmalogens and ether-lipids, resulted in reduced levels of plasmalogen and increased dyslipidemia^41^.

Interestingly, while linoleic acid (18:2; a type of omega-6 fatty acid)-containing lipids were generally underabundant, the SevO association with lipids containing arachidonic acid (20:4) acyl chains varied and did not have the same magnitude of effect. The role of arachidonic acid in adipose tissue inflammation in obesity has been reported^42^. The lower and varied directions of effect for these species suggest a minimum role for SevO in this population and warrant further research.

We also observed reduced phosphatidylcholines and phosphatidic acid species levels in individuals with SevO. Phosphatidylcholines regulate lipogenesis via sterol regulatory element-binding proteins (SREBPs), with SREBP-1 controlling genes involved in fatty acid and triglyceride synthesis, and SREBP-2 regulating cholesterol biosynthesis^43^. The concomitant decrease in phosphatidic acids may indicate disrupted phosphatidic acid phosphatase activity and impaired SREBP-mediated lipogenic signaling^44^. Alternatively, it may reflect reduced phosphatidic acid synthesis via the phospholipase D pathway, which has been implicated in obesity^45^. Further studies are needed to clarify these mechanisms.

Clinically, lipid species depleted in SevO—such as phosphatidylcholines, methylated cholesteryl esters, and sulfatides—were inversely associated with markers of inflammation, cardiac injury, liver dysfunction, insulin resistance, and adiposity. These lipids are known to constrain lipid storage (e.g., via lipid droplet regulation by phosphatidylcholines)^46^, facilitate triglyceride exchange (cholesteryl esters)^31^, and reduce inflammation (sulfatides)^47^. In contrast, lipids enriched in SevO, including diacylglycerols and triacylglycerols, were positively correlated with liver injury, insulin resistance, and adipose tissue burden. These lipids promote lipid storage, insulin resistance, inflammation^34,36^, and hormonal dysregulation (e.g., leptin and ghrelin pathways^20^). A reduced phosphatidylcholine/phosphatidylethanolamine (PC/PE) ratio in SevO further suggests impaired VLDL metabolism, compromised triglyceride transport, and hepatic lipid accumulation—hallmarks of steatosis^48^ and broader metabolic dysfunction. Given SevO’s systemic nature and strong links to CMD, these lipidomic alterations underscore potential mechanistic pathways connecting lipid metabolism, SevO, and CMD risk.

While our study offers several strengths—including a uniquely high-risk, well-characterized Hispanic/Latino cohort, a narrowly defined SevO phenotype, the largest-to-date number of species tested, and rigorous analytical methods—certain limitations must be noted. First, only one lipid species (representing a triacylglycerol with shorter length acyl chains) showed a consistent causal association with SevO, likely reflecting limitations in genetic instrument strength or the bidirectional complexity of metabolic pathways, rather than an absence of causal relationships. Nonetheless, our conservative causal approach minimized bias and highlighted the potential role of acylglycerols in SevO. Second, patterns of association with SevO were likely influenced by the chain length of each lipid species. For example, our observed inverse association between long-chain acylcarnitines and SevO did not generalize to shorter-chain species, which have shown opposite associations in the literature^49^. Similarly, discrepancies in phosphatidylserine associations may reflect differences in lipid species profiled. Third, while we focused on endogenous lipids, we also detected significant differences in exogenous, diet- or microbiota-derived lipids—such as odd-numbered and DHA-containing species—whose interpretation is complicated by emerging evidence of partial endogenous synthesis^50^. Finally, the cross-sectional design limits causal inference over time. Despite these constraints, our study provides novel insights into lipidomic mechanisms underlying SevO and establishes a critical foundation for future longitudinal and mechanistic investigations.

In summary, our integrative multi-omics analysis, leveraging clinical data alongside advanced dimensionality reduction and statistical techniques, enhances understanding the role of the lipidome in severe obesity, a major risk factor for a variety of cardiometabolic diseases. We identified statistically significant associations between SevO and a broad spectrum of lipid species and classes, several of which were predictive of SevO status and clinically relevant cardiometabolic disease (CMD) traits. Notably, triacylglycerols containing shorter length acyl chains were consistently elevated in individuals with SevO, strongly differentiated SevO versus control status, and were the standout lipid class exhibiting a consistent causal relationship with SevO. Other lipid metabolites—including ceramides, lipoproteins, acylcarnitines, phosphatidylcholines, and cholesteryl esters—also contributed to SevO differentiation, though with variable consistency. In contrast, lipids containing long and very-long acyl chains were generally depleted in SevO, suggesting modulation by population-specific nutritional and genetic factors. These findings provide novel mechanistic insights and lay the groundwork for future translational and therapeutic investigations and interventions.

## Box 1. Non-standard Abbreviations and Acronym: lipid classes assessed in this study

**Box 1. Non-standard Abbreviations and Acronym: lipid classes assessed in this study.**
| **Abr.** | **Term** | **Abr.** | **Term** |
| --- | --- | --- | --- |
| **AC** | Acylcarnitine | **LPE(P)** | Lysoalkenylphosphatidylethanolamine |
| **AC-OH** | hydroxylated acylcarnitine | **LPI** | Lysophosphatidylinositol |
| **BA** | bile acid | **methyl-CE** | Methyl-cholesteryl ester |
| **C1P** | Ceramide-1-phosphate | **methyl-DE** | Methyl-dehydrocholesteryl ester |
| **CE** | Cholesteryl ester | **OxSpecies** | Oxidised lipids |
| **Cer(d)** | Ceramide | **PA** | Phosphatidic acid |
| **Cer(m)** | Deoxyceramide | **PC** | Phosphatidylcholine |
| **COH** | Free cholesterol | **PC(O)** | Alkylphosphatidylcholine |
| **DE** | Dehydrocholesteryl ester | **PC(P)** | Alkenylphosphatidylcholine |
| **deDE** | Dehydrodesmosterol ester | **PE** | Phosphatidylethanolamine |
| **DG** | Diacylglycerol | **PE(O)** | Alkylphosphatidylethanolamine |
| **DG(O)** | Alkylmonoacylglycerol | **PE(P)** | Alkenylphosphatidylethanolamine |
| **dhCer** | Dihydroceramide | **PG** | Phosphatidylglycerol |
| **dimethyl-CE** | Dimethyl-cholesteryl ester | **PI** | Phosphatidylinositol |
| **FFA** | Free fatty-acid | **PIP1** | Phosphatidylinositol monophosphate |
| **GD1** | GD1 ganglioside | **PS** | Phosphatidylserine |
| **GM3** | GM3 ganglioside | **S1P** | Sphingosine-1-phosphate |
| **Hex2Cer** | Dihexosylceramide | **SHexCer** | Sulfatide |
| **Hex3Cer** | Trihexosylceramide | **SM** | Sphingomyelin |
| **HexCer** | Monohexosylceramide | **Sph** | Sphingosine |
| **LPC** | Lysophosphatidylcholine | **TG(NL)** | Triacylglycerol (Neutral Loss) |
| **LPC(O)** | Lysoalkylphosphatidylcholine | **TG(O)** | Alkyldiacylglycerol |
| **LPC(P)** | Lysoalkenylphosphatidylcholine | **Ubiquinone** | Ubiquinone |
| **LPE** | Lysophosphatidylethanolamine | **TG(SIM)** | Triacylglycerol (Selected Ion Monitoring) |

## Acknowledgements

The authors would like to thank the CCHC cohort team led by Rocío Uribe for recruiting and interviewing the participants; Marcela Morris, BS, and her team for laboratory support; Valley Baptist Medical Center in Brownsville, Texas, for providing space for our Center for Clinical and Translational Science Clinical Research Unit; and the community of Brownsville, along with the participants who willingly took part in this study in their city.

## Funding

This study was funded in part by Center for Clinical and Translational Sciences, National Institutes of Health Clinical and Translational Award grant no. UL1 TR000371 from the National Center for Advancing Translational Sciences, predoctoral fellowship from American Heart Association (no. 18PRE34060101 to H-H.C.), and NIH grants U01CA288325, R01HL142302, R35HG010718, R01HG011138 and R01GM140287 to E.R.G., R01HL142302, R01DK127084, R01AG078452, and R01HL163262 to J.E.B., H-H.C., K.E.N., A.G., M. L., J.B.M, and S.P.F-H. And K.E.N., P.G.-L., Y.M. A., M.G., D.K., K. L. Y. are further supported by R01HL151152, R01 DK122503, R01HD057194, R01HG010297, R01HL143885. JC and JB were supported by National Institutes of Health (NIH) grant R01 HL140681.

## Conflict of interest

The authors declare that they have no conflict of interest.

## Contributions

Conceptualization: MYA, KEN, JEB, MG. Statistical Analysis: MYA, MG, CJ, PJM, ZL. Data Curation and Visualization: MYA, HMH, Supervision: KEN, MG, JEB, SF-H, JC, JB, CJ, PJM; Resources: SF-H, JBM, MG, CJ, PJM; Writing – Original Draft: MYA, ABP, PG-L, CJ, PJM, KEN, HMH, JC, MG, JEB; Writing – Critical Review & Editing: all authors.

## Data Availability

The data supporting this study’s findings are available upon request from CCHC. The CCHC data is not publicly accessible as it contains information that could compromise participants’ privacy and consent. The administrating body can grant access to CCHC data upon a legitimate request. Summary BHS-GWAS data used in MR are available from the GWAS catalog (ebi.ac.uk/gwas/), and metabolomics.baker.edu.au.

## Notes

### Competing Interest Statement

The authors have declared no competing interest.

### Author Declarations

The CCHC study was approved by the Committee for the Protection of Human Subjects of the University of Texas Health Science Center, Houston. The written consent of participants were obtained upon inclusion in the study. The current study, titled "The Circulating Lipidome In Severe Obesity? involved no primary data collection, and used secondary data, together with summary data from publicly available repositories, and therefore the study is exempt from IRB review based on current regulations.

